# Chronic kidney risk among Nepalese migrant workers in the countries of Gulf and Malaysia: a population-based cross-sectional survey in Nepal

**DOI:** 10.1101/2025.05.15.25327703

**Authors:** Nirmal Aryal, Pramod R. Regmi, Arun Sedhain, Sankalpa Bhattarai, Radheshyam Krishna KC, Shravan Kumar Mishra, Ben Caplin, Neil Pearce, Edwin van Teijlingen

## Abstract

**Introduction:** There have been increasing concerns about possible chronic kidney disease (CKD) problems among returnee Nepalese migrant workers from the Gulf countries and Malaysia. This population-based cross-sectional survey primarily aimed to examine kidney health risks of Nepalese recent migrants compared to non-migrants from the same community.

**Methods:** We conducted a survey of 1438 participants from Dhanusha district (718 recent migrants and 720 non-migrants (including historic migrants). Recent migrants (all sexes) aged 18 years or above, who had stayed at least two years in six countries of the Gulf region or Malaysia in any occupation and had returned in the past 12 months, were included. We used the core questionnaire and protocol for the Disadvantaged Populations eGFR Epidemiology Study (DEGREE) and added questions on migration.

**Results:** All recent migrants were male and they were compared to male historic migrants, male non-migrants, and female non-migrants. Only 6 (0.4%) cases of eGFR <60mL/min/1.73m^2^ were identified overall. The prevalences of eGFR <60mL/min/1.73m^2^ were 0.4% (95% CI: 0.08 to 1.2), 0.5% (95% CI: 0.01 to 2.6), 1.2% (95% CI: 0.1 to 4.2%), and zero in male recent migrants, male historic migrants, male non-migrants, and female non-migrants respectively. In the adjusted multiple regression model, male recent migrants had a statistically non-significant, slightly reduced mean difference of −0.8 mL/min/1.73m^2^ (95%CI: −3.6 to 2.0) in eGFR compared to male non-migrants. A separate adjusted model among male recent migrants showed a strong association between reduced mean eGFR and older age, occupation as a security guard, current or past smokers, and ethnicity.

**Conclusion:** This study found a low prevalence of eGFR <60mL/min/1.73m^2^ in Nepalese recent migrants. There were no associations of mean eGFR by migration status despite male migrants being exposed to risk factors for kidney disease.

## 1. Introduction

Nearly 5 million work permits have been granted for Nepalese workers since 2008 for work-related international migration, of which the vast majority were for the Gulf Co-operation Council (GCC) countries and Malaysia ^1^. The Nepal 2021 Census reported that 23.4% of households have at least one family member living abroad ^2^. According to the Nepal Labour Migration Report 2022, labour migrants were mostly males, low-skilled, based in GCC countries and Malaysia, and had sent over US$ 8.8 billion of remittance, representing one-quarter of the national income ^1^.

In recent years, kidney health risks of Nepalese labour migrants returning from GCC countries and Malaysia have been the subject of considerable discussion by the scientific community ^3,4^, global and national media of Nepal ^5–7^, and global organisations ^89^. Although there is a paucity of evidence, some hospital-based studies indicate possible higher kidney problems among returnee Nepalese migrant workers.

A 2023 study analysing data from two large haemodialysis treatment facilities in the country reported that 28% of kidney failure patients were returnee migrants and had kidney failures at significantly lower mean age than non-migrant patients ^10^, although this may simply reflect the fact that migrants tend to be younger than non-migrants on average. A recent survey at the tertiary care centre in eastern Nepal also reported returnee migrants comprised 31% of the patients on maintenance dialysis ^11^. Another hospital-based study among 44 returnee migrant workers treated for Chronic Kidney Disease (CKD) in Nepal found that 70.5% were involved in manual or semi-manual work in GCC countries and Malaysia, 70% worked more than 60 hours per week, and the cause for CKD was unknown for 77.3% ^12^. More than two-thirds of the Nepal-based nephrologists in an online survey reported that Nepalese migrants could have a higher kidney health risk than the general Nepalese population ^3^.

The recent qualitative study among 12 returnee Nepalese migrants from GCC and Malaysia diagnosed with kidney-related problems found poor access to potable water, excessive use of pain medication, poor dietary habits, and occupational pollutants as potential risks ^13^. A longitudinal study with 65 Indian construction workers in Saudi Arabia reported that 18% suffered from kidney injury suggesting exposure to heat, long working hours, dehydration, sleep deprivation, and obesity as risk factors ^14^. A 2021 report from Amnesty International also documented that doctors in South Asia noted high levels of kidney health-related issues in returnee migrants from GCC countries and Malaysia ^8^.

Nepal does not have a national disease surveillance system, nor do hospitals keep records by migration status. Thus, the health condition of returnee migrants is not recorded unless they claim financial compensation for any of the 15 critical illnesses (including kidney failure). Regular health screening of migrant workers in GCC countries is not legally bound, rather sporadic, and thus usually impossible to detect incremental health risks. Therefore, it is difficult to ascertain whether a significant number of Nepalese returnee migrants with kidney failures and kidney health problems seeking treatment in Nepal’s hospitals reflect a similar risk in the general population or whether this is an indication of a disproportionately higher risk related to them being abroad.

A possible higher risk among Nepalese migrants should not be overlooked because they are young (median age 28, with half being between 25 and 34 years) ^1^. Furthermore, all aspiring migrants have to clear mandatory pre-migration health screening in Nepal which also includes serum creatinine test for kidney function. In addition, the working and living conditions of many Nepalese migrants in GCC countries and Malaysia include known risk factors for kidney health, such as exposure to searing heat, heavy workload, dehydration, excessive use of alcohol, sugary drinks, and non-steroidal anti-inflammatory drugs (NSAIDS) ^13,15^. These factors, individually or in combination, can affect kidney function.

The primary aim of this study is to generate the first-ever population-based evidence on kidney health risks and concomitant risk factors in returnee migrant workers and to compare them to non-migrant populations in Nepal in the district with the highest transnational outmigration.

## 2. Methods

This was a survey of the whole source population with previous work-related migration history as a primary exposure of interest. Data were collected in mid-2023. Dhanusha district in the east of the country was selected as it has the highest number of labour out-migrants in Nepal ^1^, with the vast majority being low-skilled (e.g., construction and factory) workers. Potential participants were invited if they were: (i) recent migrants (all sexes) aged 18 years and over; had stayed at least two years (to allow for exposure to migrant worker experience) in GCC countries or Malaysia in any occupation and had returned in the past 12 months, or (ii) non-migrants or migrants who had returned >12 months ago (all sexes) aged 18-50 years; who lived in the study area for at least two years, and (iii) speak and understand Nepali language. Those who were unable to provide written or verbal consent, pregnant, or with severe impairments were excluded.

Considering the 5% prevalence of eGFR <60mL/min/1.73m^2^ in the non-migrant population from the likely age group in Nepal ^16^ and assuming eGFR <60mL/min/1.73m^2^ prevalence twice as likely as in the non-migrant population (10%), we estimated 435 samples in each migrant and non-migrant group (i.e., 870 in total) yielding 80% power and 5% margin of error. While adjusting it with a 1.5 design effect and 10% non-response rate, we required 1,436 samples in total (718 in each group).

### 2.1 Sampling technique

A multistage sampling method was applied. As the sampling frame on migrants by administrative units was unavailable, the study team consulted with related stakeholders and local authorities to identify the urban or rural municipalities in Dhanusha district with the highest number of out-migrants. Accordingly, recruitment was focused on Laximiniya (rural) and Chhireshowrnath (urban) municipalities. In each municipality, information on recent migrants was collected and approached to confirm eligibility criteria.

Those meeting the inclusion criteria from their households were invited. For households without migrants, only one participant meeting inclusion criteria out of every seven households was invited. This was based on our mapping exercise which estimated that 12.5% of households in the district had at least one returnee migrant. The process continued until the required number of participants was recruited, which consisted of a total of 16 wards (lowest administrative units) from two municipalities. A study flow diagram detailing study areas and participants is presented in Figure 1.

**Figure 1:**
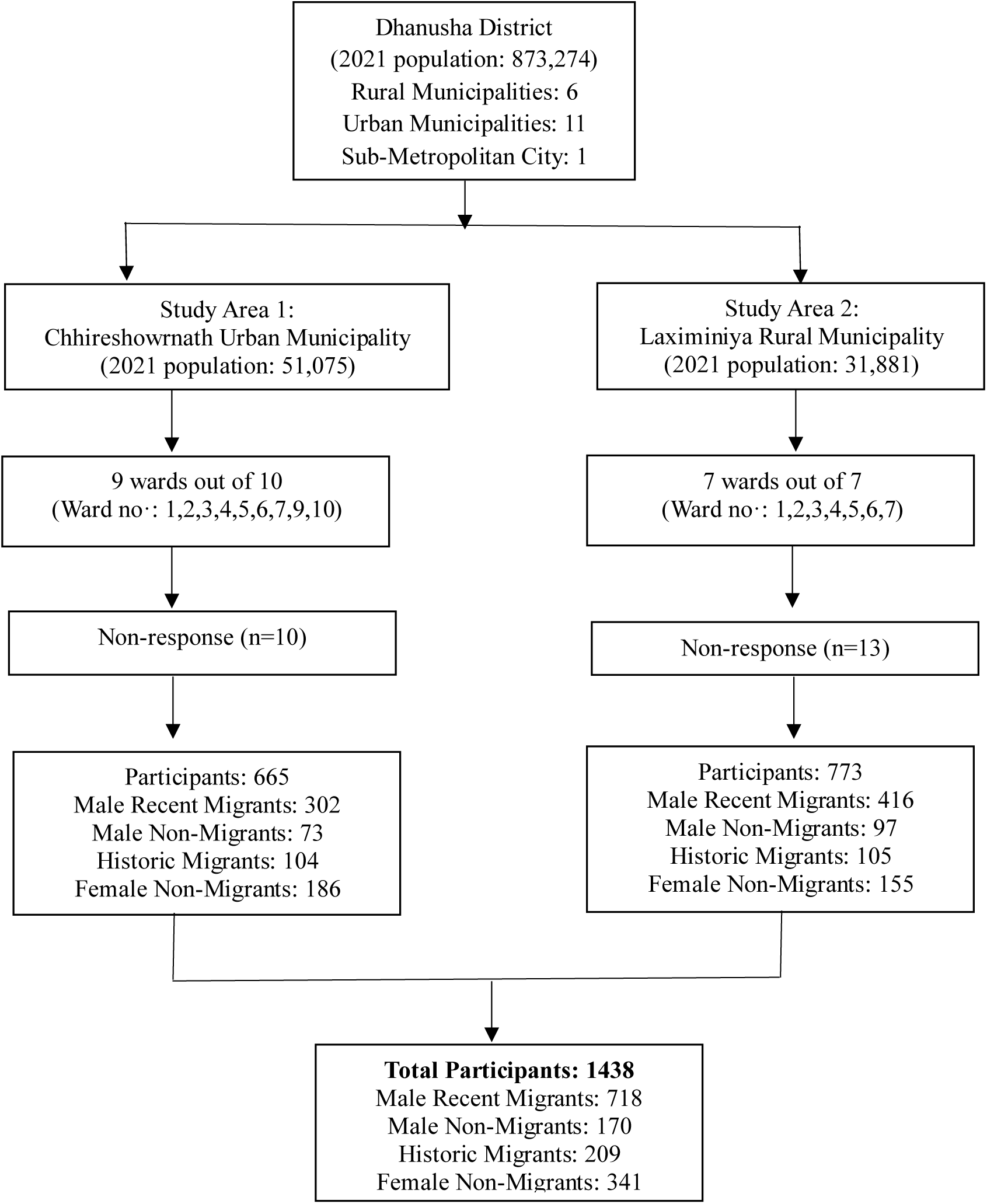
Study areas and sampling frame segregated by participant number and migration status

### 2.2 Data collection tool and operational definitions

A survey tool was developed based on the core questionnaire for the Disadvantaged Populations eGFR Epidemiology Study (DEGREE) ^17^ with additional questions on migration history, living and working conditions while abroad, and kidney health-related risk factors (e.g. exposure to heat, water intake, chemical exposure). The questionnaire was translated into Nepali and pre-tested with five returnee migrants to ensure clarity, intelligibility, and cultural sensitivity.

Returnee migrants as per the inclusion criteria are defined as recent or historic migrants. The eGFR was calculated using the 2021 CKD-EPI equation ^18,19^. A low eGFR suggestive of underlying CKDu was defined by excluding common known causes of CKD, i.e, the absence of diabetes mellitus, hypertension, or urinary evidence of glomerulonephritis in the form of heavy proteinuria ^17,20^.

Sex was defined as assigned at birth. Blood pressure was classified as hypertensive if mean systolic or diastolic blood pressure was equal to or greater than 140 mmHg or 90 mmHg, respectively, or if participants were on antihypertensive medication. Participants were classified as living with diabetes if their fasting or non-fasting blood glucose level was equal to or greater than 126 mg/dL or 200 mg/dL, respectively or if they were on antidiabetic medication. Proteinuria and heavy proteinuria were defined as high if the urinary albumin to creatinine ratio (ACR) was greater than 30 mg/g and equal to or greater than 300mg/g, respectively. Overweight or obesity was defined by body mass index (BMI) equal to or greater than 25 kg/m^2^.

### 2.3 Data collection techniques

Two laboratory technicians and five research assistants (RAs) with public health/nursing degrees were employed. Data was collected in the morning as participants were likely to be at work during the day. If absent, up to three attempts were made before the next eligible participant on the list was approached. The RAs completed a paper version of the questionnaire. Laboratory technicians collected blood (5 mL) and urine samples at the same time. Blood samples were stored at 2-8^0^ C using a cold pack and transported to the laboratory the same day. We took a sample from each batch and recorded the batch variation. The project coordinator supervised the field data collection, monitored data quality, and dealt with any problems at the field level. A repeat test of serum creatinine and proteinuria was planned after 3 months among those participants with eGFR <90 ml/min/1.73m^2^ or urinary ACR greater than 30 mg/g. Serum creatinine assays were referenced according to the Isotope Dilution Mass Spectrometry (IDMS) standard.

With consent from the participants, the RAs reviewed medical records if available and recorded relevant information. These records were requested directly from the participants because there was no other way to access them.

The participants were provided with a participant information sheet in Nepali with detailed information about the study and what the participation would involve. Written consent was obtained where possible; however, witnessed consent was obtained if participants were illiterate. Participants with abnormal results were also referred to the nearest hospital with a nephrologist.

Ethical approvals were provided by Bournemouth University (UK) (reference number: 44154, approval date: 19 July 2022)) and the Nepal Health Research Council (reference number: 262, approval date: 11 August 2022). The STROBE statement was followed for reporting the results.

### 2.4 Statistical analysis

Descriptive statistics including the Chi-squared test and Student’s T-test (or its non-parametric counterpart Mann-Whitney test) were used to compare categorical (e.g., hypertension, diabetes) and continuous outcomes (e.g., eGFR, proteinuria), respectively. Multiple linear regressions were used to (i) estimate the association between mean eGFR and migration status, and (ii) factors associated with reduced eGFR among recent migrants adjusted with the potential confounding variable (age) and known risk factors. Assumptions were checked for linear regressions which were reasonably met. Stata software (Version 17) was used for analysis. Among historic migrants, only one was female and thus excluded from data analysis for male-to-male comparisons.

We could not access a significant number of eligible participants during the repeat tests after three months, particularly for recent migrants (45.2% for reduced eGFR and 65.4% for high proteinuria were missing), and hence only the results of the first blood and urine tests were included here.

### 2.5 Role of the funding source

The funder does not have any role in any aspect of the study.

### 2.6 Patient and public involvement and engagement (PPIE)

We held consultation meetings with key stakeholders working for migrant communities, researchers, health care providers and migrant workers in Malaysia and Nepal to ensure that the issue of kidney health risks among Nepalese migrants is significant and warrants further investigation ^21^. Members of the research advisory group, including returnee migrants and organisations focused on migration health in Nepal, contributed to the formulation of research questions, the exploration of various study designs considering time and resources, the translation of study tools, the piloting of the study questionnaire, and the dissemination of study findings.

## 3. Results

Twenty-three potential participants (1.6%) refused to take part citing reasons such as fear of needles and possible effects on their migration plans. A total of 718 male recent migrants, 209 male historic migrants, 170 male non-migrants, and 342 female non-migrants participated. The 209 male historic migrants had mainly worked in the six countries of the Gulf and Malaysia (94.7%) but had returned at least one year before.

Key socio-demographic characteristics, lifestyle risk factors, biological risk factors, and medical history of the study participants are shown in Table 1. The mean age of male recent migrants and male historic migrants was the same (35.0 years) which was significantly higher than other groups. Around two-thirds of male recent migrants, male historic migrants, male non-migrants, and more than half of the female non-migrants were from similar ethnic groups (‘castes’). Male recent migrants reported a higher monthly mean income than rest of the groups (including all sources and from family sharing the same kitchen), and the mean difference among the groups was statistically significant.

**Table 1:**
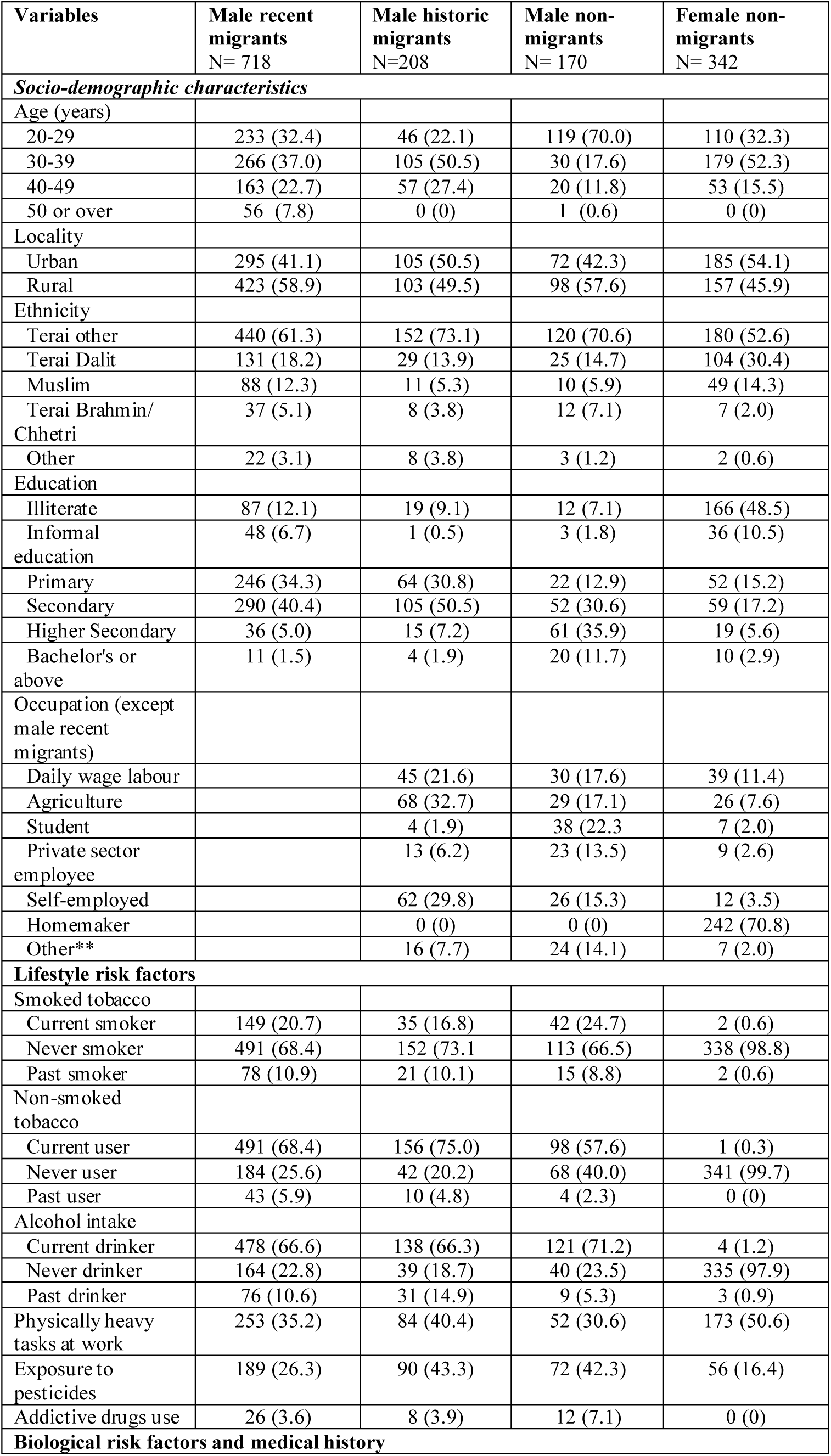
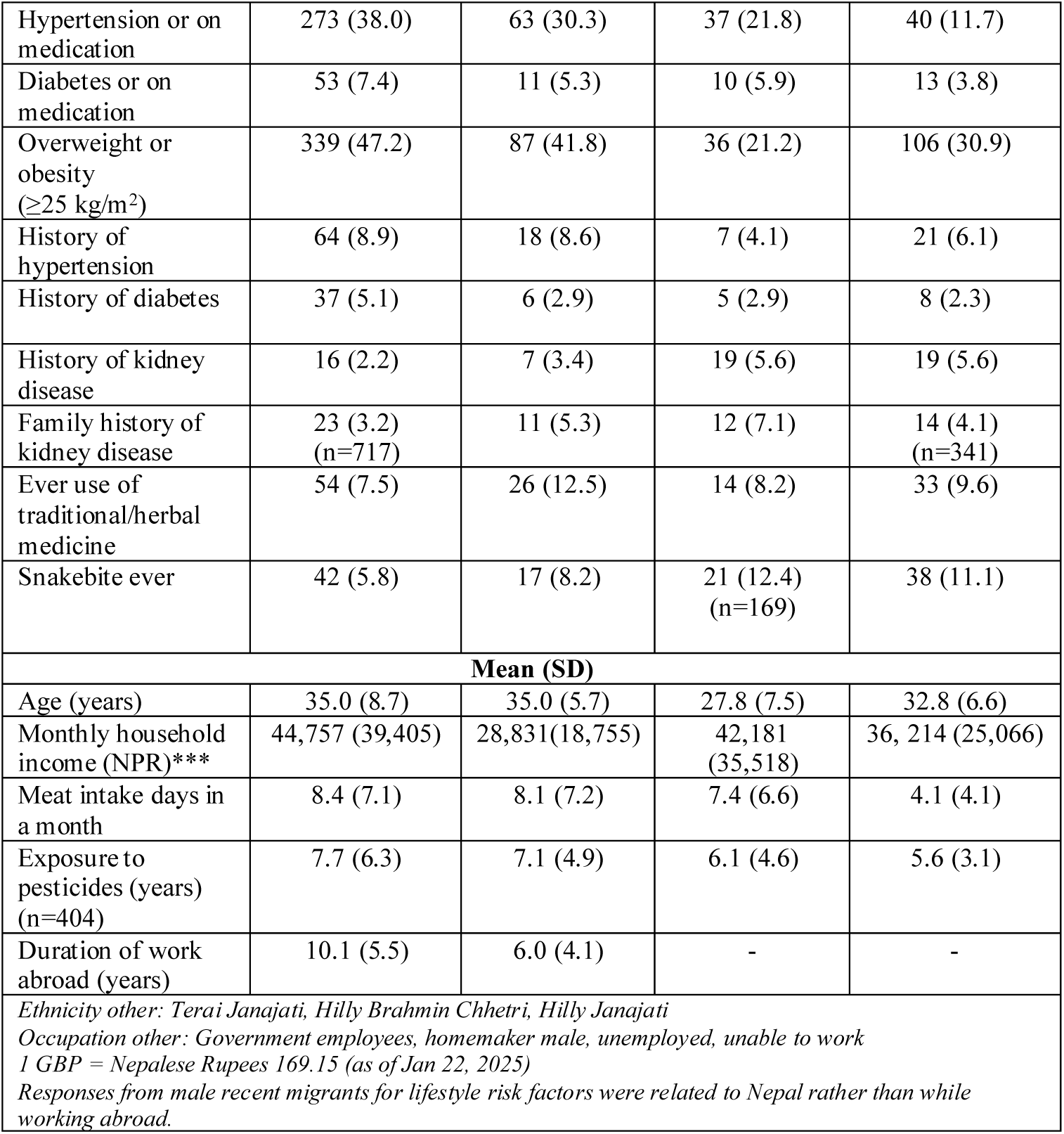
Key socio-demographic characteristics, lifestyle, biological risk factors, and medical history.

| Variables | Male recent migrants<br>N= 718 | Male historic migrants<br>N=208 | Male non-migrants<br>N= 170 | Female non-migrants<br>N= 342 |
| --- | --- | --- | --- | --- |
| ***Socio-demographic characteristics*** |  |  |  |  |
| Age (years) |  |  |  |  |
| 20-29 | 233 (32.4) | 46 (22.1) | 119 (70.0) | 110 (32.3) |
| 30-39 | 266 (37.0) | 105 (50.5) | 30 (17.6) | 179 (52.3) |
| 40-49 | 163 (22.7) | 57 (27.4) | 20 (11.8) | 53 (15.5) |
| 50 or over | 56 (7.8) | 0 (0) | 1 (0.6) | 0 (0) |
| Locality |  |  |  |  |
| Urban | 295 (41.1) | 105 (50.5) | 72 (42.3) | 185 (54.1) |
| Rural | 423 (58.9) | 103 (49.5) | 98 (57.6) | 157 (45.9) |
| Ethnicity |  |  |  |  |
| Terai other | 440 (61.3) | 152 (73.1) | 120 (70.6) | 180 (52.6) |
| Terai Dalit | 131 (18.2) | 29 (13.9) | 25 (14.7) | 104 (30.4) |
| Muslim | 88 (12.3) | 11 (5.3) | 10 (5.9) | 49 (14.3) |
| Terai Brahmin/<br>Chhetri | 37 (5.1) | 8 (3.8) | 12 (7.1) | 7 (2.0) |
| Other | 22 (3.1) | 8 (3.8) | 3 (1.2) | 2 (0.6) |
| Education |  |  |  |  |
| Illiterate | 87 (12.1) | 19 (9.1) | 12 (7.1) | 166 (48.5) |
| Informal education | 48 (6.7) | 1 (0.5) | 3 (1.8) | 36 (10.5) |
| Primary | 246 (34.3) | 64 (30.8) | 22 (12.9) | 52 (15.2) |
| Secondary | 290 (40.4) | 105 (50.5) | 52 (30.6) | 59 (17.2) |
| Higher Secondary | 36 (5.0) | 15 (7.2) | 61 (35.9) | 19 (5.6) |
| Bachelor's or above | 11 (1.5) | 4 (1.9) | 20 (11.7) | 10 (2.9) |
| Occupation (except male recent migrants) |  |  |  |  |
| Daily wage labour |  | 45 (21.6) | 30 (17.6) | 39 (11.4) |
| Agriculture |  | 68 (32.7) | 29 (17.1) | 26 (7.6) |
| Student |  | 4 (1.9) | 38 (22.3) | 7 (2.0) |
| Private sector employee |  | 13 (6.2) | 23 (13.5) | 9 (2.6) |
| Self-employed |  | 62 (29.8) | 26 (15.3) | 12 (3.5) |
| Homemaker |  | 0 (0) | 0 (0) | 242 (70.8) |
| Other** |  | 16 (7.7) | 24 (14.1) | 7 (2.0) |
| ***Lifestyle risk factors*** |  |  |  |  |
| Smoked tobacco |  |  |  |  |
| Current smoker | 149 (20.7) | 35 (16.8) | 42 (24.7) | 2 (0.6) |
| Never smoker | 491 (68.4) | 152 (73.1) | 113 (66.5) | 338 (98.8) |
| Past smoker | 78 (10.9) | 21 (10.1) | 15 (8.8) | 2 (0.6) |
| Non-smoked tobacco |  |  |  |  |
| Current user | 491 (68.4) | 156 (75.0) | 98 (57.6) | 1 (0.3) |
| Never user | 184 (25.6) | 42 (20.2) | 68 (40.0) | 341 (99.7) |
| Past user | 43 (5.9) | 10 (4.8) | 4 (2.3) | 0 (0) |
| Alcohol intake |  |  |  |  |
| Current drinker | 478 (66.6) | 138 (66.3) | 121 (71.2) | 4 (1.2) |
| Never drinker | 164 (22.8) | 39 (18.7) | 40 (23.5) | 335 (97.9) |
| Past drinker | 76 (10.6) | 31 (14.9) | 9 (5.3) | 3 (0.9) |
| Physically heavy tasks at work | 253 (35.2) | 84 (40.4) | 52 (30.6) | 173 (50.6) |
| Exposure to pesticides | 189 (26.3) | 90 (43.3) | 72 (42.3) | 56 (16.4) |
| Addictive drugs use | 26 (3.6) | 8 (3.9) | 12 (7.1) | 0 (0) |
| ***Biological risk factors and medical history*** |  |  |  |  |
| Hypertension or on medication | 273 (38.0) | 63 (30.3) | 37 (21.8) | 40 (11.7) |
| Diabetes or on medication | 53 (7.4) | 11 (5.3) | 10 (5.9) | 13 (3.8) |
| Overweight or obesity ($\geq 25$ kg/m<sup>2</sup>) | 339 (47.2) | 87 (41.8) | 36 (21.2) | 106 (30.9) |
| History of hypertension | 64 (8.9) | 18 (8.6) | 7 (4.1) | 21 (6.1) |
| History of diabetes | 37 (5.1) | 6 (2.9) | 5 (2.9) | 8 (2.3) |
| History of kidney disease | 16 (2.2) | 7 (3.4) | 19 (5.6) | 19 (5.6) |
| Family history of kidney disease | 23 (3.2)<br>(n=717) | 11 (5.3) | 12 (7.1) | 14 (4.1)<br>(n=341) |
| Ever use of traditional/herbal medicine | 54 (7.5) | 26 (12.5) | 14 (8.2) | 33 (9.6) |
| Snakebite ever | 42 (5.8) | 17 (8.2) | 21 (12.4)<br>(n=169) | 38 (11.1) |
| **Mean (SD)** |  |  |  |  |
| Age (years) | 35.0 (8.7) | 35.0 (5.7) | 27.8 (7.5) | 32.8 (6.6) |
| Monthly household income (NPR)*** | 44,757 (39,405) | 28,831(18,755) | 42,181<br>(35,518) | 36, 214 (25,066) |
| Meat intake days in a month | 8.4 (7.1) | 8.1 (7.2) | 7.4 (6.6) | 4.1 (4.1) |
| Exposure to pesticides (years) (n=404) | 7.7 (6.3) | 7.1 (4.9) | 6.1 (4.6) | 5.6 (3.1) |
| Duration of work abroad (years) | 10.1 (5.5) | 6.0 (4.1) | - | - |
| *Ethnicity other: Terai Janajati, Hilly Brahmin Chhetri, Hilly Janajati*<br>*Occupation other: Government employees, homemaker male, unemployed, unable to work*<br>*1 GBP = Nepalese Rupees 169.15 (as of Jan 22, 2025)*<br>*Responses from male recent migrants for lifestyle risk factors were related to Nepal rather than while working abroad.* |  |  |  |  |

Non-migrant males had a higher prevalence of current smoking and current alcohol intake compared to other groups, and male historic migrants were more likely to be current non-smoked tobacco users. The differences in proportions for these variables were statistically significant. The most popular smokeless tobacco used by all groups was chewing plain tobacco leaves or mixed with betel nuts, and beer was the most common alcohol consumed. The prevalence of these risk factors among female non-migrants was negligible. Supplementary statistics S1 shows the bivariate analyses of the variables included in Table 1.

Male recent migrants worked in the GCC countries and Malaysia; more than half had worked in Qatar (Supplementary Figure S2), had worked abroad for an average of about ten years, and 54.6% were repeat migrants. More than two-thirds were involved in factory and construction work (Supplementary Figure S3).

Half of the male recent migrants were allowed just once or no rest time during the day and on average took 2.4 (SD 0.1) days off a month (Supplementary Table S4). One-quarter of jobs involved physically heavy work and nearly half (45.5%) considered heat exposure level at work to be high. More than one-third (38.3%) were always exposed to dust at work. Nearly 10% were always exposed to chemicals and just 1.8% were always exposed to pesticides. Very few migrants used painkillers regularly. The overwhelming majority reported easy access to water and toilets at work. Half of the migrants (53.5%) reported drinking alcohol while abroad and around 40% drank one standard drink of alcohol at least 2-3 times per week. Beer was the most common type of alcohol for 80% of migrants, and 15.3% also drank counterfeit or homebrewed alcohol.

Compared to other groups, a higher proportion of male recent migrants were hypertensive (or on medication), diabetic (or on medication), and overweight or obese. After adjustment for age, male recent migrants and male historic migrants had significantly higher odds for overweight or obesity [Odds Ratio (OR) for male recent migrants: 2.15, 95% CI: 1.49 to 3.09; OR for male historic migrants: 1.56, 95% CI: 1.00 to 2.41] compared with male non-migrants. No such associations were observed for hypertension (or on medication) and diabetes (or on medication) after adjusting for age.

Overall, 49 participants (3.4%) had a history of kidney disease which was significantly higher in male and female non-migrants (5.6% in each group) compared to male recent and historic migrants. Of these, the vast majority had kidney stones (n=38, 77.5%), particularly among non-migrant females (n=15, 30.6%) and male recent migrants (n=12, 24.5%). A total of 60 participants (4.2%, n=1436) reported having a family history of kidney disease, with a significantly higher proportion among non-migrant males compared to other groups. A minority of participants (8.8%) had ever used traditional/herbal medicine, mainly for gastritis issues, and this use was particularly higher for male historic migrants.

Six participants (0.4%) (95% CI: 0.1 to 0.9) had eGFR <60 mL/min/1.73m^2^ with no statistically difference in proportions between participant groups (P=0.28) (Table 2). The eGFR <60mL/min/1.73m^2^ prevalence was 0.4% (95% CI: 0.08 to 1.2) among male recent migrants, 0.5% (95% CI: 0.01 to 2.6) among male historic migrants, highest among male non-migrants (1.2%) (95% CI: 0.1 to 4.2), and zero among female non-migrant. Of these, two-thirds (4 out of 6, 66.7%) were from the age group 30 to 39 years. The age-adjusted prevalence of eGFR <60mL/min/1.73m^2^ was 0.2% for male recent migrants, 0.2% for male historic migrants, and 1.2% for male non-migrants. Figure 2 presents histograms of eGFR distribution by participants’ groups. A higher proportion of female non-migrants had increased levels of proteinuria (>30 mg/g) at 10.5% (95% CI: 7.5 to 14.3) compared to other groups but this difference was statistically not significant (P=0.45).

**Figure 2:**
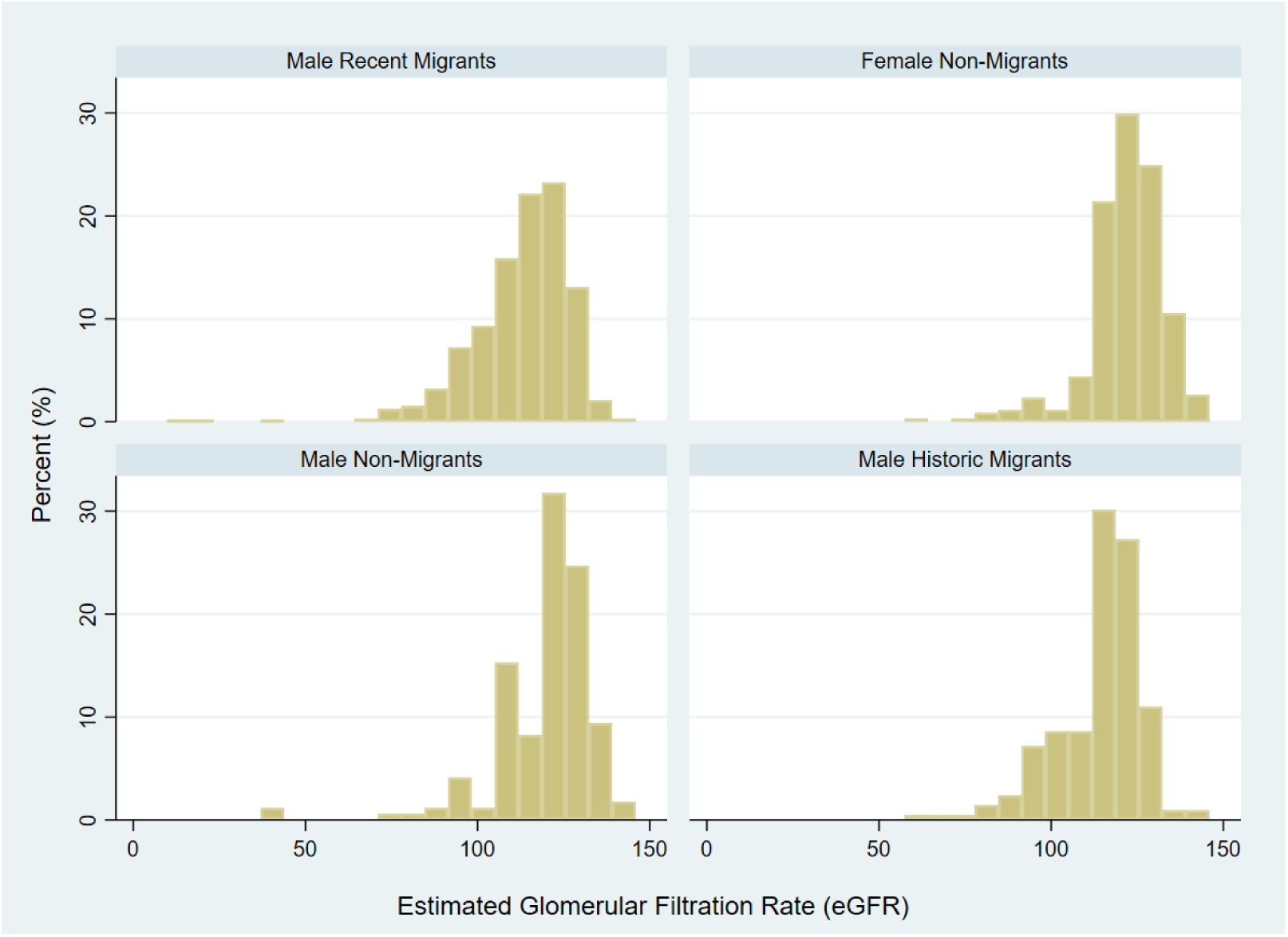
Distribution of estimated Glomerular Filtration Rate (eGFR) by migration status

**Table 2:** Prevalence of reduced eGFR and high proteinuria.

| Variables | Male recent migrants<br>N= 718<br>(n,%) (95% CI) | Male historic migrants<br>N=208<br>(n,%) (95% CI) | Male non-migrants<br>N= 170<br>(n,%) (95% CI) | Non-migrant females<br>N= 342<br>(n,%) (95% CI) |
| --- | --- | --- | --- | --- |
| eGFR<60 mL/min/1.73m<sup>2</sup> | 3 (0.4)<br>(0.08 to 1.2) | 1 (0.5)<br>(0.01 to 2.6) | 2 (1.2)<br>(0.1 to 4.2) | 0 (0) |
| eGFR <60 mL/min/1.73m<sup>2</sup> without hypertension, diabetes, heavy proteinuria* | 0 (0) | 0 (0) | 1 (0.6)<br>(0.01 to 3.2) | 0 (0) |
| eGFR <90 mL/min/1.73m<sup>2</sup> | 42 (5.8)<br>(4.2 to 7.8) | 11 (5.3)<br>(2.6 to 9.2) | 6 (3.5)<br>(1.3 to 7.5) | 9 (2.6)<br>(1.2 to 4.9) |
| eGFR <90 mL/min/1.73m<sup>2</sup> without hypertension, diabetes, heavy proteinuria* | 13 (1.8)<br>(3.2 to 9.9) | 6 (2.9)<br>(1.1 to 6.1) | 5 (2.9)<br>(35.9 to 99.6) | 8 (2.3)<br>(0.9 to 6.7) |
| Proteinuria >30 mg/g | 55 (7.7)<br>(5.8 to 9.8) | 20 (9.6)<br>(5.9 to 14.4) | 15 (8.8)<br>(5.0 to 14.1) | 36 (10.5)<br>(7.5 to 14.3) |
| **Mean (SD)** |  |  |  |  |
| eGFR (mL/min/1.73m<sup>2</sup>) | 112.8 (14.0)<br>(111.8 to 113.8) | 114.0 (12.6)<br>(112.3 to 115.7) | 119.7 (14.5)<br>(117.5 to 121.9) | 122.0 (11.2)<br>(120.8 to 123.2) |
| Albumin to Creatinine ratio (mg/g) | 13.2 (51.4)<br>(9.4 to 17.0) | 21.7 (97.9)<br>(8.4 to 35.3) | 27.8 (214.1)<br>(-4.6 to 60.2) | 17.3 (57.8)<br>(11.2 to 23.5) |
| Serum creatinine (mg/dL) | 0.9 (0.3)<br>(0.85 to 0.89) | 0.8 (0.1)<br>(0.83 to 0.87) | 0.8 (0.2)<br>(0.82 to 0.88) | 0.6 (0.1)<br>(0.57 to 0.59) |
| *Heavy proteinuria: Albumin to Creatinine ratio ≥300 mg/g |  |  |  |  |

Multiple linear regression estimated the association between mean eGFR and migration status adjusted for potential confounding variables and known risk factors (Table 3). In the adjusted model, the mean difference (95% CI) in eGFR among male recent migrants compared to male non-migrants was −0.8 mL/min/1.73m^2^ (−3.6 to 2.0) with a wider confidence interval (P=0.58). Female non-migrants had significantly higher mean eGFR difference (95% CI) compared to male non-migrants; 4.1 mL/min/1.73m^2^ (1.2 to 7.1), P=0.006. Higher age, history of kidney disease, current or past smokers and ethnicity were associated with the lower eGFR. Among ethnic groups, the Terai Dalit group had a significantly lower mean eGFR when compared to the reference group, the Terai other. Sensitivity analyses were performed excluding outlier observations in eGFR, age and BMI, which yielded a similar result, except for the negative association between being current or past drinkers and mean eGFR.

**Table 3:**
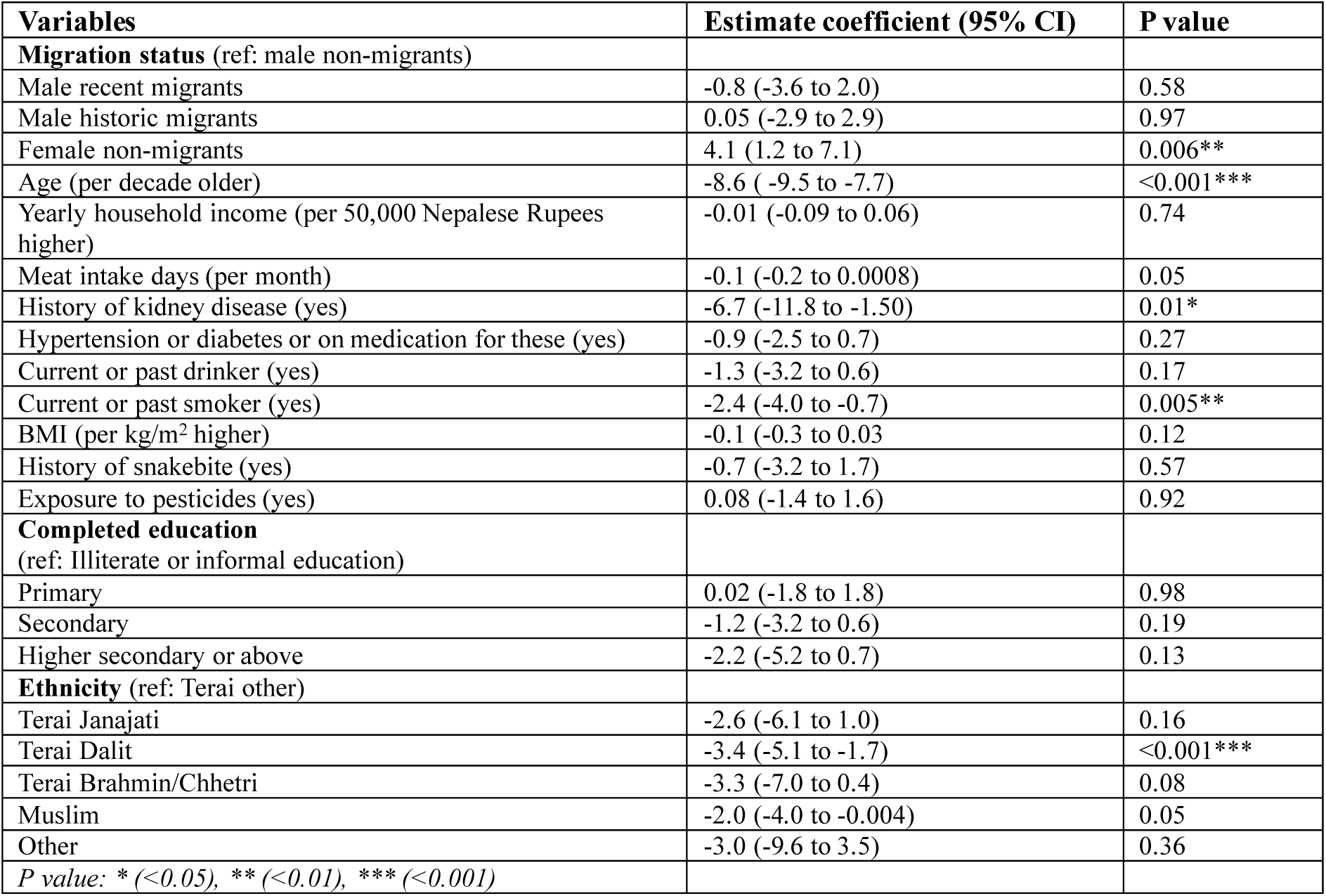
Multiple regression model for the association between mean eGFR and migration status adjusted for potential confounding variables and known risk factors.

| **Variables** | **Estimate coefficient (95% CI)** | **P value** |
| --- | --- | --- |
| **Migration status** (ref: male non-migrants) |  |  |
| Male recent migrants | -0.8 (-3.6 to 2.0) | 0.58 |
| Male historic migrants | 0.05 (-2.9 to 2.9) | 0.97 |
| Female non-migrants | 4.1 (1.2 to 7.1) | 0.006** |
| Age (per decade older) | -8.6 (-9.5 to -7.7) | <0.001*** |
| Yearly household income (per 50,000 Nepalese Rupees higher) | -0.01 (-0.09 to 0.06) | 0.74 |
| Meat intake days (per month) | -0.1 (-0.2 to 0.0008) | 0.05 |
| History of kidney disease (yes) | -6.7 (-11.8 to -1.50) | 0.01* |
| Hypertension or diabetes or on medication for these (yes) | -0.9 (-2.5 to 0.7) | 0.27 |
| Current or past drinker (yes) | -1.3 (-3.2 to 0.6) | 0.17 |
| Current or past smoker (yes) | -2.4 (-4.0 to -0.7) | 0.005** |
| BMI (per kg/m<sup>2</sup> higher) | -0.1 (-0.3 to 0.03) | 0.12 |
| History of snakebite (yes) | -0.7 (-3.2 to 1.7) | 0.57 |
| Exposure to pesticides (yes) | 0.08 (-1.4 to 1.6) | 0.92 |
| **Completed education** (ref: Illiterate or informal education) |  |  |
| Primary | 0.02 (-1.8 to 1.8) | 0.98 |
| Secondary | -1.2 (-3.2 to 0.6) | 0.19 |
| Higher secondary or above | -2.2 (-5.2 to 0.7) | 0.13 |
| **Ethnicity** (ref: Terai other) |  |  |
| Terai Janajati | -2.6 (-6.1 to 1.0) | 0.16 |
| Terai Dalit | -3.4 (-5.1 to -1.7) | <0.001*** |
| Terai Brahmin/Chhetri | -3.3 (-7.0 to 0.4) | 0.08 |
| Muslim | -2.0 (-4.0 to -0.004) | 0.05 |
| Other | -3.0 (-9.6 to 3.5) | 0.36 |
| *P value: \*(<0.05), \*\*(<0.01), \*\*\*(<0.001)* |  |  |

Table 4 shows the multiple linear regression among male recent migrants identifying the variables associated with the reduced eGFR. Higher age, occupation as a security guard, being from Terai Dalit and Muslim ethnic groups, and being current or past smokers were independently associated with the reduced eGFR. Work-related circumstances speculated to be the potential causes of kidney health risks had a negative relationship with the eGFR; however, the differences in risks were not statistically significant. No notable differences were observed while conducting sensitivity analysis excluding outlier observations in age, BMI, eGFR, and migration years. Another multiple regression analysis excluding occupational mediators from this model yielded similar results except statistically significant negative associations between BMI and eGFR (Supplementary Table S6).

**Table 4:** Factors associated with the reduced mean eGFR among male recent migrants adjusted with potential confounding variables and known risk factors.

| Variables | Estimate coefficient (95% CI) | P value |
| --- | --- | --- |
| Migration duration (years) | 0.02 (-0.2 to 0.2) | 0.87 |
| Age (per decade older) | -7.3 (-8.7 to -5.8) | <0.001*** |
| Yearly household income (per 50,000 Nepalese Rupees higher) | 0.02 (-0.1 to 0.1) | 0.65 |
| Number of days off in a month | 0.01 (-0.5 to 0.5) | 0.95 |
| Meat intake days (per month) | -0.04 (-0.2 to 0.1) | 0.52 |
| **Occupation (ref: other<sup>#</sup>)** |  |  |
| Construction work | -2.7 (-5.7 to 0.1) | 0.06 |
| Factory work | -1.4 (-4.0 to 1.3) | 0.31 |
| Driving | -1.4 (-4.9 to 2.2) | 0.45 |
| Security guard | -9.0 (-15.1 to -3.0) | 0.003** |
| **Physical intensity at work (ref: none or low)** |  |  |
| Moderate | -0.05 (-2.8 to 2.7) | 0.97 |
| High | -0.7 (-3.2 to 1.9) | 0.59 |
| **Heat exposure at work (ref: none or low)** |  |  |
| Moderate | -0.5 (-3.5 to 2.4) | 0.72 |
| High | 1.1 (-2.1 to 4.3) | 0.49 |
| **Work setting (ref: indoor)** |  |  |
| Indoor and indoor mix | -0.8 (-3.2 to 1.6) | 0.49 |
| Outdoor | 0.6 (-1.8 to 3.1) | 0.60 |
| **Dust exposure at work (always or sometimes compared to never or rarely)** | -0.6 (-3.2 to 1.9) | 0.64 |
| History of kidney disease (yes) | -9.0 (-20.1 to 2.0) | 0.11 |
| Hypertension or diabetes or on medication (yes) | -1.6 (-3.9 to 0.8) | 0.19 |
| Current or past drinker (yes) | -1.4 (-4.0 to 1.2) | 0.29 |
| Current or past smoker (yes) | -2.3 (-4.3 to -0.3) | 0.02* |
| BMI (per kg/m<sup>2</sup> higher) | -0.2 (-0.5 to 0.01) | 0.06 |
| **Completed education (ref: Illiterate or informal education)** |  |  |
| Primary | 0.3 (-2.6 to 3.3) | 0.82 |
| Secondary | -0.7 (-3.5 to 2.2) | 0.65 |
| Higher secondary or above | -3.4 (-10.3 to 3.5) | 0.33 |
| **Ethnicity (ref: Terai other)** |  |  |
| Terai Janajati | -1.5 (-5.7 to 2.7) | 0.49 |
| Terai Dalit | -5.3 (-7.9 to -2.6) | <0.001*** |
| Terai Brahmin/Chhetri | 0.4 (-3.4 to 4.3) | 0.83 |
| Muslim | -3.1 (-6.1 to -0.1) | 0.04* |

P value: \* (<0.05), \*\* (<0.01), \*\*\* (<0.001); Other<sup>#</sup>= mainly office work, sales, hotel work, cleaning

## 4. Discussion

This study found only 6 cases (0.4%) of eGFR <60mL/min/1.73m^2^. The crude and age-adjusted prevalence of eGFR <60mL/min/1.73m^2^ was higher among male non-migrants compared to male recent migrants, male historic migrants and female non-migrants. Two-thirds of these cases were reported among the younger age group (30 and 39 years). Just one participant (male non-migrant) had CKDu (eGFR<60mL/min/1.73m^2^ in the absence of diabetes, hypertension or heavy proteinuria).

In the adjusted model, male recent migrants had a lower mean eGFR compared to male non-migrants, however, this was clinically insignificant with a wide confidence interval. Male historic migrants and female non-migrants showed a higher mean eGFR compared to male non-migrants; this was statistically significant among female non-migrants only. A separate multiple regression among a sub-group of male recent migrants showed a strong association between a lower mean eGFR and occupation as a security guard.

The present study’s findings are similar to studies comparing Ghanaians in Europe and Ghanaians in Ghana, which showed a higher cardiometabolic risk but lower onset and occurrence of CKD among migrants ^22,23^. The only nationally representative study in Nepal reported a prevalence of CKD among 4.4% of people aged 20 to 59 years old ^24^, however, it used a different equation than ours for calculating eGFR and defined CKD as eGFR <60 mL/min/1.73m^2^ and/or albumin to creatinine ratio of ≥30 mg/g. When compared to this national study and using the same criteria, our study reported a prevalence of 8.8% among all participant groups. The difference of at least five years in data collection points between the national study and the present study may indicate that the burden of kidney health problems is increasing rapidly at the community level.

In the present study, an association between reduced mean eGFR and occupation as a security guard of recent migrants is of note. Security guards reported poorer work-related circumstances than those in other occupations, these included the longest daily working hours, the lowest daily rest time, the lowest number of days off in a month, and the highest proportion with poor access to toilets. We could not identify any previously published quantitative evidence on these work-related conditions; however, our previous engagement with Nepalese migrant workers in Malaysia identified these issues among Nepalese security guards ^21^.

As expected, work-related circumstances of male recent migrants, such as high heat exposure, outdoor work, and physically heavy work, were associated with a quantitatively lower eGFR, but the associations were statistically non-significant.

The present study found a significantly higher prevalence of hypertension, diabetes, and overweight/obesity in male recent migrants and male historic migrants compared to male and female non-migrants. Compared to male recent migrants and male historic migrants, the latest World Health Organization (WHO) STEPS survey 2019 reported lower prevalences of these risk factors in the general adult population in Nepal aged between 15 and 69 years old, whilst these were similar to non-migrant males and females in the present study ^25^. Interestingly, risk factors such as alcohol intake and smoking habits were more prevalent in non-migrant males. However, 15.3% of male recent migrants reported consuming potentially hazardous counterfeit or home-brewed alcohol while working abroad. Thus, both poor lifestyle habits and work-related circumstances may have contributed to cardiometabolic risks in male recent migrants while abroad.

The study also reported an independent association between reduced eGFR and participants from Terai Dalit and Muslim populations in separate adjusted models for both all participants and male recent migrants only. These are socially and economically one of Nepal’s most disadvantaged and underserved population groups. A nationwide study also documented an association between Terai Dalit ethnicity and eGFR <60mL/min/1.73m^2^ ^24^. In the present study, participants from these ethnicities usually did physically demanding jobs; non-migrants from these groups were mostly daily wage labourers and recent migrants were security guards and construction workers. High proteinuria was observed more frequently among female non-migrants. A further investigation is warranted to ascertain the causes of high proteinuria in the young female population.

Despite the absence of a high burden of kidney disease, this study has further underpinned the precarious working conditions of Nepalese migrants in GCC and Malaysia (such as long working hours with one or no break time, unavailability of indoor rest place during the break, not taking weekly days off regularly, and regular exposure to heat) which could have negative health impacts ^15^.

### Strengths and limitations

This is the first population-based study globally that compared kidney health risks among migrants (from GCC countries and Malaysia) and non-migrants from the same community. This is also the largest study so far on Nepalese migrants documenting work-related and lifestyle risks while working abroad. The main limitation is that we did not include the results of the repeat test (after three months) due to the low turnout rate. We could not access enough eligible participants during the repeat tests, particularly male recent migrants because they usually have two months of leave every two to three years and they had already gone abroad again for work. We also selected study areas purposively to achieve the required sample size of migrants. Although a sample of migrants is random, a random sampling of the whole population was not possible because of the logistics and time constraints to recruit enough migrants. Finally, we do not know if CKD-EPI creatinine equation accurately estimates the actual eGFR in Nepalese populations. Although this might have implications on the absolute number with eGFR <60mL/min/1.73m^2^, it does not impact the conclusion regarding differences between the groups.

In conclusion, this population-based study has found a low prevalence of eGFR <60mL/min/1.73m^2^ and eGFR <60mL/min/1.73m^2^ in the absence of evidence for known causes of CKD in both migrant and non-migrant populations in Nepal. In particular, male recent migrants were not at higher risk of eGFR <60mL/min/1.73m^2^ compared to non-migrant males, despite being exposed to risky work conditions and lifestyle-related factors. However, this study indicates that specific sub-groups of Nepalese migrants working in GCC and Malaysia, mainly those working as security guards and from Terai Dalit and Muslim ethnicities, could be at higher risk. These require further investigations. Known risk factors of kidney and cardiometabolic health risks, mainly hypertension, diabetes, and overweight/obesity were found to be significantly higher among recent and historic migrants compared to non-migrant males and females. Thus, this study also implies possible future kidney health risks among Nepalese migrants. Although this is a cross-sectional study, it has added new information to this much-discussed but evidence-scarce area.

## Data Availability

The dataset of this study is available from the corresponding author upon reasonable request.

## Disclosures

Nothing to disclose

## Data Sharing

The dataset of this study is available from the corresponding author upon reasonable request.

## Acknowledgements

We would like to thank the field research team, our participants, and stakeholders in the study area for their support. A special thanks goes to our collaborators in Nepal, namely Green Tara Nepal, the International Organisation for Migration (IOM) in Kathmandu, and the Madhes Pradesh Public Health Laboratory, for their collaboration and logistical support in Nepal. Finally, we are grateful to the Colt Foundation (in the UK) for the financial support for this study.

## Supplementary Materials

*Supplementary Statistics S1*: Bivariate analyses of key socio-demographic characteristics, lifestyle, biological risk factors, and medical history of participants categorised into four groups (male recent migrants, male historic migrants, male non-migrants, and female non-migrants) (Microsoft Word Document)

*Supplementary Figure S2*: Destination countries of current migrants (N=718) (Microsoft Excel)

*Supplementary Figure S3*: Occupation of recent migrants during their recent employment (Microsoft Excel)

*Supplementary Table S4*: Working conditions and lifestyle characteristics of male recent migrants (N=718) (Microsoft Word Document)

*Supplementary Table S5*: Factors associated with reduced mean eGFR among male recent migrants adjusted for confounding factors, known risk factors and excluding occupational mediators (Microsoft Word Document)

*Supplementary STROBE Statement*

## Notes

### Competing Interest Statement

The authors have declared no competing interest.

### Funding Statement

The Colt Foundation, UK, funded this study.

### Author Declarations

Ethical approvals were provided by Bournemouth University Research Ethics Committee (BU REC), UK (reference number: 44154, approval date: 19 July 2022)) and the Nepal Health Research Council Ethical Review Board (reference number: 262, approval date: 11 August 2022).

