## Supplementary Materials for "Chronic kidney risk among Nepalese migrant workers in the countries of Gulf and Malaysia: a population-based cross-sectional survey in Nepal"

**Supplementary Statistics S1: Bivariate analyses of key socio-demographic characteristics, lifestyle, biological risk factors, and medical history of participants categorised into four groups (male recent migrants, male historic migrants, male non-migrants, and female non-migrants)**

*1. Categorical variables (Chi-squared test)*

A· Socio-demographic characteristics

(i) Age-groups: Pearson $X^2$ (9, N=1438)=182·4, P <0·001

(ii) Locality: Pearson $X^2$ (3, N=1438) = 18·5, P <0·001

(iii) Ethnicity: Pearson $X^2$ (18, N=1438) = 519·7, P <0·001

B· Lifestyle risk factors

(i) Smoked tobacco: Pearson $X^2$ (6, N=1438)=132·7, P <0·001

(ii) Non-smoked tobacco: Pearson $X^2$ (6, N=1438)=580·7, P <0·001

(iii) Alcohol intake: Pearson $X^2$ (6, N=1438)=634·7, P <0·001

(iv) Physically heavy tasks at work: Pearson $X^2$ (3, N=1438)=28·8, P <0·001

(v) Exposure to pesticides: Pearson $X^2$ (3, N=1438)=64·9, P <0·001

(vi) Addictive drugs use: Pearson $X^2$ (9, N=1436)=56·7, P <0·001

C· Biological risk factors and medical history

(i) Hypertension or on medication: Pearson $X^2$ (3, N=1438)=83·0, P <0·001

(ii) Diabetes or on medication: Pearson $X^2$ (3, N=1438)=5·5, P=0·14

(iii) Overweight or obesity: Pearson $X^2$ (3, N=1438)=52·6, P <0·001

(iv) History of hypertension: Pearson $X^2$ (3, N=1438)=6·0, P=0·11

(v) History of kidney disease: Pearson $X^2$ (6, N=1438)=81·7, P <0·001

(vi) Family history of kidney disease: Pearson $X^2$ (6, N=1436)=90·5, P <0·001

(vii) Ever use of traditional/herbal medicine: Pearson $X^2$ (3, N=1438)=5·4, P <0·001

(viii) Snakebite ever: Pearson $X^2$ (6, N=1437)=20·7, P=0·002

*2. Mean variables (one-way ANOVA/Kruskal-Wallis test)*

(i) Age: F (3, 1434) = 43·6, P <0·001

(ii) Monthly income: F (3, 1410)= 14·0, P<0·001

(iii) Meat intake days in a month: F (3, 1433)= 35·2 P<0·001

(iv) Exposure to pesticides (years): Kruskal-Wallis [Chi-squared]= 3·3, P=0·34

**Supplementary Figure S2: Destination countries of current migrants (N=718)**

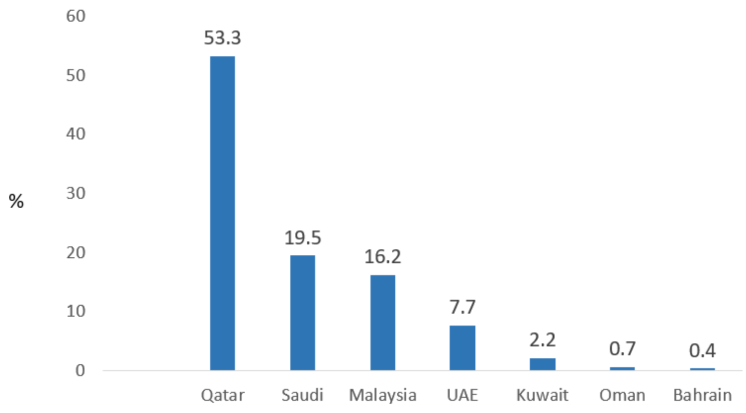

**Supplementary Figure S3: Occupation of recent migrants during their recent employment**

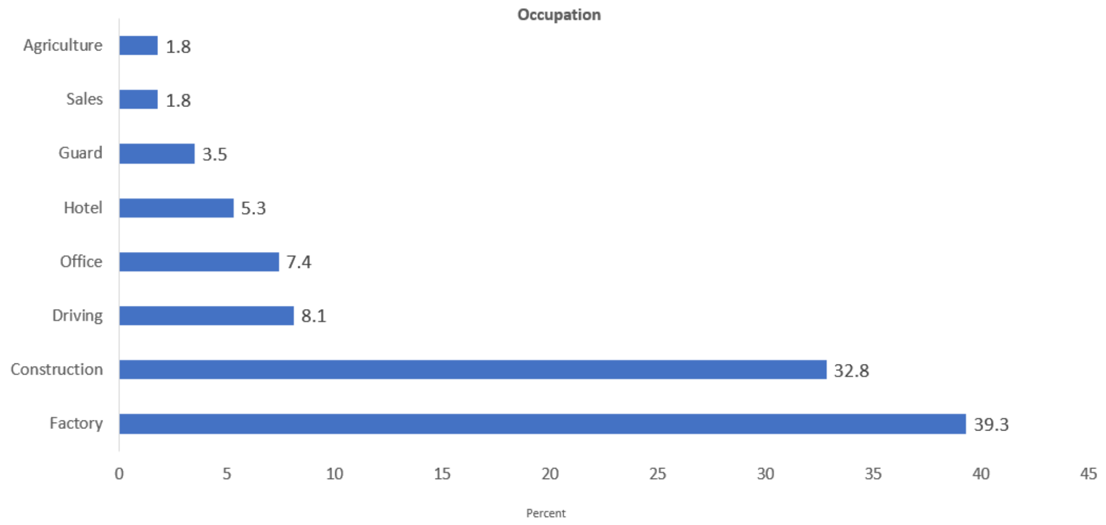

**Supplementary Table S4: Working conditions and lifestyle characteristics of male recent migrants (N=718)**

| Variables | Number (%) | 95% CI |
|---|---|---|
| **Rest episode(s) at work/day (n=717)** | | |
| One time or less | 365 (50.9) | 47.1 to 54.6 |
| Two times | 233 (32.5) | 29.1 to 36.1 |
| Three times | 89 (12.4) | 10.1 to 15.0 |
| Four times or more | 30 (4.2) | 2.8 to 5.9 |
| **Rest place at work** | | |
| Inside workplace | 272 (37.9) | 34.3 to 41.5 |
| Hostel or accommodation | 230 (19.2) | 28.6 to 35.6 |
| Shade | 138 (19.2) | 16.4 to 22.3 |
| Outside workplace | 39 (5.4) | 3.9 to 7.3 |
| Other (e.g., vehicle, canteen) | 39 (5.4) | 3.9 to 7.3 |
| **Regular day off once a week** | | |
| Always | 392 (54.6) | 50.9 to 58.3 |
| Sometimes | 244 (44.0) | 30.5 to 37.6 |
| Never | 82 (11.4) | 9.2 to 13.9 |
| **Physical level at work** | | |
| Heavy | 185 (25.8) | 22.6 to 29.1 |
| Moderate | 271 (37.7) | 34.2 to 41.4 |
| Light | 262 (36.5) | 32.9 to 40.1 |
| **Work setting** | | |
| Indoor | 332 (46.2) | 42.5 to 49.9 |
| Outdoor | 213 (29.7) | 26.3 to 33.1 |
| Indoor and outdoor mix | 173 (24.1) | 21.0 to 27.4 |
| **Heat exposure level at work** | | |
| High | 327 (45.5) | 41.8 to 49.3 |
| Moderate | 145 (20.2) | 17.3 to 23.3 |
| Low or none | 246 (34.3) | 30.8 to 37.9 |
| **Water availability at work** | 700 (97.5) | 96.1 to 98.5 |
| **Easy toilet access at work** | 697 (97.1) | 95.6 to 98.2 |
| **Dust exposure at work** | | |
| Always | 275 (38.3) | 34.8 to 41.9 |
| Sometimes | 120 (16.7) | 14.0 to 19.6 |
| Rarely or never | 323 (45.0) | 41.3 to 48.7 |
| **Chemical exposure at work** | | |
| Always | 69 (9.6) | 7.5 to 12.0 |
| Sometimes | 71 (9.9) | 7.8 to 12.3 |
| Rarely or never | 578 (80.5) | 77.4 to 83.3 |
| **Pesticide exposure at work** | | |
| Always or sometimes | 13 (1.8) | 0.9 to 3.1 |
| Never or rarely | 705 (98.2) | 96.9 to 99.0 |
| **Painkiller use** | | |
| Minimum once a week | 29 (4.0) | 2.7 to 5.7 |
| More than once a week | 22 (3.1) | 1.9 to 4.6 |
| Sometimes | 491 (68.4) | 64.8 to 71.8 |
| Never | 176 (24.5) | 21.4 to 27.8 |
| **Alcohol intake while abroad (yes)** | 384 (53.5) | 49.7 to 58.2 |
| **Frequency consuming one standard drink of alcohol (n=384)** | | |
| Daily | 7 (1.8) | 0.7 to 3.7 |
| 3-4 days a week | 18 (4.7) | 2.8 to 7.3 |
| 1-2 days a week | 134 (34.9) | 30.1 to 39.9 |
| 1-3 days a month | 148 (38.5) | 33.6 to 43.6 |
| Less than once a month | 77 (20.0) | 16.2 to 24.4 |
| | **Mean (SD)** | |
| Average working hours/day | 10.7 (0.1) | 10.5 to 10.8 |
| Average rest duration/day (minutes) | 76.7 (1.3) | 74.2 to 79.2 |
| Average no of days off in a month | 2.4 (0.1) | 2.2 to 2.5 |

**Supplementary Table S5: Factors associated with reduced mean eGFR among male recent migrants adjusted for confounding factors, known risk factors and excluding occupational mediators**

| Variables | Estimate coefficient (95% CI) | P value |
|---|---|---|
| Migration duration (years) | 0.02 (-0.2 to 0.2) | 0.89 |
| Age (per decade older) | -7.4 (-8.8 to -5.9) | <0.001*** |
| Yearly household income (per 50,000 Nepalese Rupees higher) | 0.01 (-0.1 to 0.1) | 0.71 |
| Meat intake days (per month) | -0.03 (-0.1 to 0.1) | 0.58 |
| History of kidney disease (yes) | -8.9 (-19.8 to 2.1) | 0.10 |
| Hypertension or diabetes or on medication (yes) | -1.5 (-3.9 to 0.9) | 0.22 |
| Current or past drinker (yes) | -1.1 (-3.6 to 1.3) | 0.37 |
| Current or past smoker (yes) | -2.3 (-4.2 to -0.3) | 0.02* |
| BMI (per $kg/m^2$ higher) | -0.2 (-0.5 to -0.003) | 0.04* |
| **Occupation (ref: other#)** | | |
| Construction work | -2.5 (-5.1 to 0.1) | 0.06 |
| Factory work | -1.4 (-4.0 to 1.3) | 0.32 |
| Driving | -1.3 (-4.7 to 2.1) | 0.44 |
| Security guard | -8.8 (-14.6 to -3.1) | 0.003** |
| **Completed education** (ref: Illiterate or informal education) | | |
| Primary | 0.2 (-2.8 to 3.1) | 0.91 |
| Secondary | -0.8 (-3.6 to 1.9) | 0.54 |
| Higher secondary or above | -3.5 (-9.9 to 2.8) | 0.27 |
| **Ethnicity (ref: Terai other)** | | |
| Terai Janajati | -1.4 (-5.6 to 2.8) | 0.51 |
| Terai Dalit | -5.3 (-7.9 to -2.7) | <0.001*** |
| Terai Brahmin/Chhetri | 0.1 (-3.8 to 4.1) | 0.95 |
| Muslim | -3.1 (-6.1 to -0.1) | 0.04* |
| *P value: * (<0.05), ** (<0.01), *** (<0.001); Other# = mainly office work, sales, hotel work, cleaning* | | |

STROBE Statement—Checklist of items that should be included in reports of ***cross-sectional studies***

| | Item No | Recommendation | Page No |
|---|---|---|---|
| **Title and abstract** | 1 | (*a*) Indicate the study's design with a commonly used term in the title or the abstract | 1 |
| | | (*b*) Provide in the abstract an informative and balanced summary of what was done and what was found | 1 |
| **Introduction** | | | |
| Background/rationale | 2 | Explain the scientific background and rationale for the investigation being reported | 2-3 |
| Objectives | 3 | State specific objectives, including any prespecified hypotheses | 3 |
| **Methods** | | | |
| Study design | 4 | Present key elements of study design early in the paper | 4 |
| Setting | 5 | Describe the setting, locations, and relevant dates, including periods of recruitment, exposure, follow-up, and data collection | 4,5 |
| Participants | 6 | (*a*) Give the eligibility criteria, and the sources and methods of selection of participants | 4, 5 |
| Variables | 7 | Clearly define all outcomes, exposures, predictors, potential confounders, and effect modifiers. Give diagnostic criteria, if applicable | 5 |
| Data sources/ measurement | 8* | For each variable of interest, give sources of data and details of methods of assessment (measurement). Describe comparability of assessment methods if there is more than one group | 5 |
| Bias | 9 | Describe any efforts to address potential sources of bias | 4-7 |
| Study size | 10 | Explain how the study size was arrived at | 4 |
| Quantitative variables | 11 | Explain how quantitative variables were handled in the analyses. If applicable, describe which groupings were chosen and why | 6-7 |
| Statistical methods | 12 | (*a*) Describe all statistical methods, including those used to control for confounding | 6-7 |
| | | (*b*) Describe any methods used to examine subgroups and interactions | Not Applicable |
| | | (*c*) Explain how missing data were addressed | 7 |
| | | (*d*) If applicable, describe analytical methods taking account of sampling strategy | Not Applicable |
| | | (*e*) Describe any sensitivity analyses | 12,13 |

| | | | |
|---|---|---|---|
| **Results** | | | |
| Participants | 13* | (a) Report numbers of individuals at each stage of study—eg numbers potentially eligible, examined for eligibility, confirmed eligible, included in the study, completing follow-up, and analysed | 8 |
| | | (b) Give reasons for non-participation at each stage | 8 |
| | | (c) Consider use of a flow diagram | 5 |
| Descriptive data | 14* | (a) Give characteristics of study participants (eg demographic, clinical, social) and information on exposures and potential confounders | 8-10 |
| | | (b) Indicate number of participants with missing data for each variable of interest | 8-10 |
| Outcome data | 15* | Report numbers of outcome events or summary measures | 11 |
| Main results | 16 | (*a*) Give unadjusted estimates and, if applicable, confounder-adjusted estimates and their precision (eg, 95% confidence interval). Make clear which confounders were adjusted for and why they were included | 11-14 |
| | | (*b*) Report category boundaries when continuous variables were categorized | 12 |
| | | (*c*) If relevant, consider translating estimates of relative risk into absolute risk for a meaningful time period | Not Applicable |
| Other analyses | 17 | Report other analyses done—eg analyses of subgroups and interactions, and sensitivity analyses | 13-14 |
| **Discussion** | | | |
| Key results | 18 | Summarise key results with reference to study objectives | 15 |
| Limitations | 19 | Discuss limitations of the study, taking into account sources of potential bias or imprecision. Discuss both direction and magnitude of any potential bias | 17 |
| Interpretation | 20 | Give a cautious overall interpretation of results considering objectives, limitations, multiplicity of analyses, results from similar studies, and other relevant evidence | 17, 18 |
| Generalisability | 21 | Discuss the generalisability (external validity) of the study results | 17 |
| **Other information** | | | |
| Funding | 22 | Give the source of funding and the role of the funders for the present study and, if applicable, for the original study on which the present article is based | 7,18 |
